# Joint Longitudinal Hazard Modeling Distinguishes Polygenic Effects on Dementia via Premorbid Cognitive Function and Aging-Related Decline

**DOI:** 10.64898/2026.11.06.26367441

**Authors:** Margaret L. Clapp Sullivan, Javier de la Fuente, Janie Corley, Sarah E. Harris, Ian J. Deary, Tom C. Russ, Simon R. Cox, Elliot M. Tucker-Drob

## Abstract

Alzheimer’s disease and related dementias (AD/ADRD) are clinical disorders characterized by declines in cognitive function to levels at which everyday living is meaningfully impaired. Genetic variants may confer risk for AD/ADRD by way of steeper rates of aging-related cognitive declines or their association with lower premorbid levels of cognitive function that place individuals closer to a clinically severe threshold of cognitive function. We applied joint longitudinal-hazard models to 26-year data from an ancestrally diverse US sample of N = 11,518 adults to estimate effects of multi-ancestry polygenic indices (PGIs) for a range of established risk factors on dementia hazard and the degree they operate via premorbid cognitive function and longitudinal cognitive decline. *APOE* e4 and PGIs for ADRD, type 2 diabetes, stroke, LDL cholesterol, and hearing loss operated via rate of change, whereas PGIs for educational attainment, cognitive function, physical activity, BMI, smoking, loneliness, schizophrenia, and externalizing were associated with dementia hazard via premorbid differences in cognitive function. Some results varied by continental ancestry. Our implementation of joint longitudinal-hazard models allowed us to distinguish how different genetic risk factors for AD/ADRD operate via degenerative processes that might be therapeutically targeted during later life and via premorbid levels of cognitive function that would require intervention earlier in the life course.

## Introduction

Alzheimer’s Disease and Alzheimer’s Disease Related Dementias (AD/ADRD) are major neurocognitive disorders of aging whose core clinical feature is cognitive deterioration that interferes with independence in everyday activities (*Diagnostic and Statistical Manual of Mental Disorders*, 2022). Recent estimates of the global prevalence of AD/ADRD indicate increases from approximately 1% at age 65 years to upwards of 30% at age 90 years (Nichols et al., 2022). However, these statistics grossly underestimate the prevalence of neurocognitive changes underlying AD/ADRD, as prodromal neurodegeneration occurs long before individuals meet clinical criteria for mild cognitive impairment (MCI) or AD/ADRD (Jack et al., 2018). As put by the National Institute on Aging-Alzheimer’s Association workgroups, “the pathophysiological process of [AD/ADRD] begins years, if not decades, before the diagnosis of clinical dementia” (Sperling et al., 2011). Indeed, aging-related cognitive declines in the general population of apparently healthy adults are strongly linked to declines in everyday functions and eventual clinical AD/ADRD diagnosis (Tucker-Drob, 2011, 2019).

Genetic risk for AD/ADRD is characterized by moderately large effects of common variants within the APOE region, highly penetrant rare variants, and a more distributed polygenic background (Andrews et al., 2023; de la Fuente et al., 2022; Scheltens et al., 2021). Several risk factors for AD/ADRD have been established in the epidemiological literature (Livingston et al., 2024), many of which are known to be heritable, and may therefore contribute to genetic risk for AD/ADRD, either through vertical pleiotropy (in which genetic effects are transmitted through a causal chain from the risk factor to AD/ADRD) or horizontal pleiotropy (in which the genetic effect on the risk factor and AD/ADRD operate through overlapping biological pathways). Either form of pleiotropy, however, may confer risk for dementia either via effects on neurodegenerative process that affect rate of cognitive decline from an earlier level of functioning, or premorbid process that affect the attained level of cognitive function prior to decline. This distinction is critical for understanding AD/ADRD etiology and determining targets for both therapeutics and public health policy. Because AD/ADRD is clinically diagnosed when an individual’s cognitive function has declined to a threshold below which their adaptive behaviors are meaningfully impaired, both genetic and epidemiological risk factors for AD/ADRD can operate via associations on premorbid levels of cognitive function that place individuals closer to the threshold prior to experiencing normal decline without those risk factors being associated with steeper rate of decline (Lövdén et al., 2020; Tucker-Drob, 2019). Processes affecting neurodegeneration and premorbid cognitive function may each be valuable targets for prevention efforts, the latter likely requiring interventions at earlier points in the life course (Walhovd et al., 2023).

Our analyses capitalize on genetic, cognitive, and epidemiological data from the Health and Retirement Study (HRS), a prospective, multiethnic longitudinal study of over 30,000 middle-aged and older US adults assessed on cognitive function approximately every 2 years for up to 25 years and prospectively ascertained for incident dementia. We compute polygenic indices (PGIs) from independent multi-ancestry GWAS discovery data corresponding to modifiable risk factors identified in the 2024 Lancet Commission on Dementia report (Livingston et al., 2024) along with a selection of other genetic predictors implicated in AD/ADRD. To directly estimate the extent to which a given risk factor for dementia hazard operates via rate of cognitive decline vs. premorbid cognitive function, we employ joint longitudinal-hazard models that integrate a growth curve approach to modeling cognitive change over time with a hazard modeling approach to predict timing of dementia onset. The advantages of the HRS in terms of sample size, representativeness, and longitudinal span are tempered by the lack of in-person assessments. Therefore, to establish the validity of inferences based on the HRS, we first present data confirming strong agreement between results from analyses of educational attainment and *APOE* genotype (two of the most well-established risk factors for AD/ADRD) in the HRS to results from the Lothian Birth Cohort of 1936 (LBC1936), a prospective longitudinal study of cognitive aging with gold standard cognitive assessments and clinical ascertainment of AD/ADRD. We then report results from the large-scale HRS for an expanded range of genetic and epidemiological risk factors.

## Results

### Overview of joint longitudinal-hazard modeling

We modelled trajectories of change in cognitive function in relation to dementia hazard using joint longitudinal-hazard models (**Online Methods).** The joint longitudinal-hazard model integrates a longitudinal mixed model, which decomposes repeated measures of cognitive function into a component capturing variation in baseline level and a component capturing subsequent rate of longitudinal cognitive change, with a hazard model of dementia onset. By incorporating a time-invariant predictor (measured at or prior to baseline) into the model, the joint model allows us to estimate the extent to which a predictor’s association with dementia hazard acts via effects on baseline cognitive level and longitudinal rate of cognitive change. We employed this approach to estimate the extent to which a given risk factor for dementia operated via premorbid adult cognitive function (cognitive level) and prodromal cognitive decline (cognitive slope). To estimate how rates of prodromal cognitive change related to subsequent dementia onset, cognitive data from timepoints concurrent with or after dementia onset were set to missing for all analyses.

### Risk for dementia onset via Child Cognitive Function, Educational Attainment, APOE, and Comparison across the LBC1936 and HRS

The HRS cohort is a nationally representative, highly powered study with expansive survey data (N = 30,509; age 51+ at baseline and 65+ for full cognitive measurements (see **Online Methods** and **Supplementary Note**); 13 waves of assessment spanning a total of 26 years; Sonnega et al., 2014). However, the survey nature of the study necessitates brief, remotely administered, cognitive assessments and algorithmically-(as opposed to clinician-) determined ascertainment of dementia. To establish the validity of inferences based on the HRS, we conducted preliminary longitudinal-hazard analyses for two well-established predictors of dementia, educational attainment and number of *APOE* e4 alleles. We then compare these HRS-based results to results of analyses of the LBC1936, a prospective longitudinal study of cognitive aging with gold-standard cognitive assessments and clinical ascertainment of dementia (N=989 participants; age 70 years at baseline; 5 waves of assessment spanning a total of 12.54 years; Corley et al., 2023; Mullin et al., 2023; Taylor et al., 2018). LBC1936 also has a high-quality measure of child cognitive function assessed at 11 years, which allows us to confirm that variation in child cognitive function is related to dementia hazard via growth curve level (baseline adult cognitive function) (Scottish Council for Research in Education, 1933). Despite being well-positioned for these validation purposes, the LBC1936 is too small to test the broader set of predictors that we seek to test in the larger HRS sample and, unlike the HRS, it is neither diverse nor nationally representative. We first report results in the HRS European-like (EUR) ancestry subsample for more direct comparisons to the Lothian Birth Cohort (Figure 1, Table S1), with results for the full, ancestrally heterogenous HRS sample and other ancestry subsamples presented in Figures 3 and 4 (Table S2-S8).

**Figure 1.**
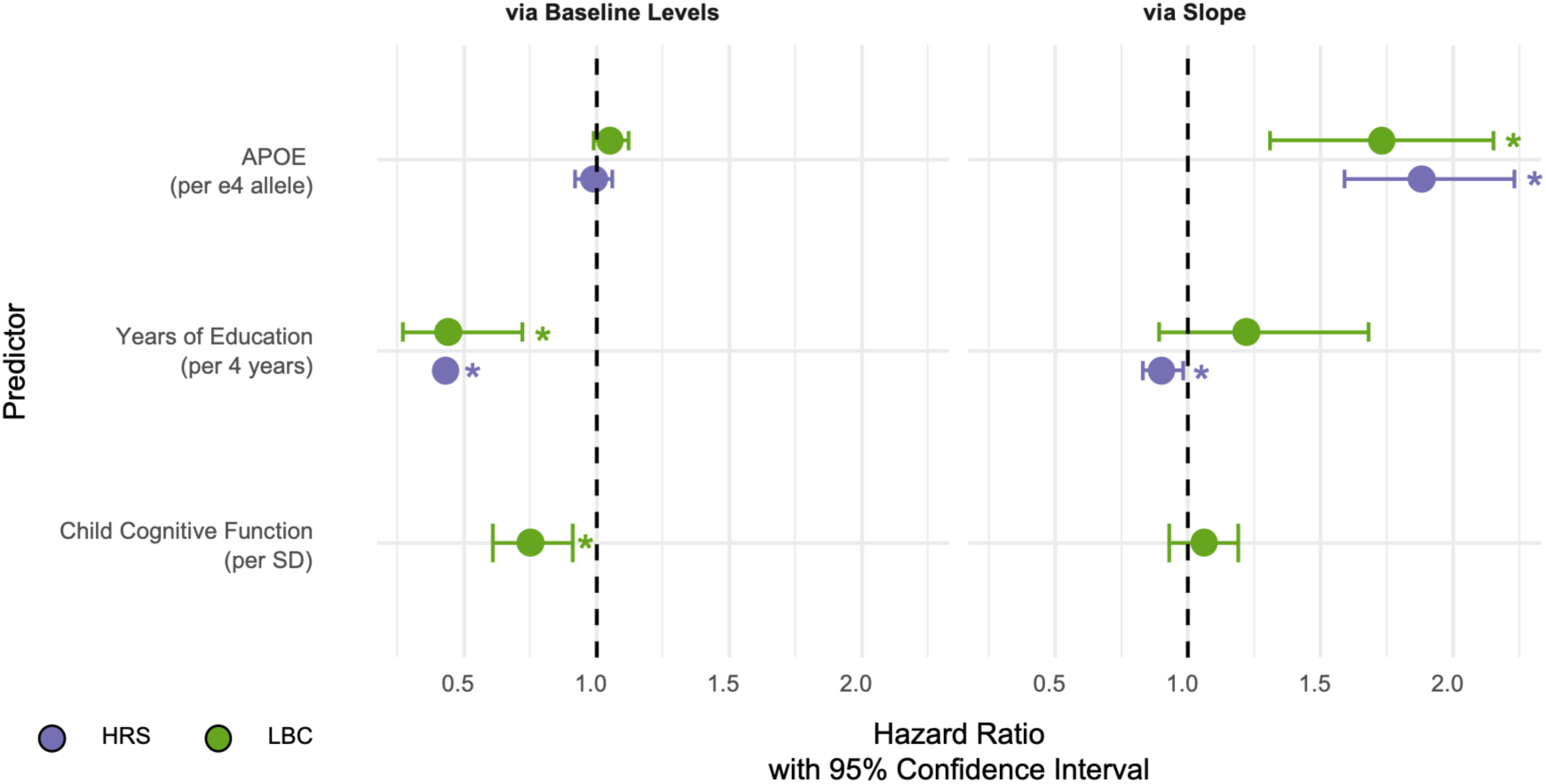
Comparison of hazard ratios in LBC and HRS for confirmatory predictors via baseline levels (left) and change (right) pathways. Estimates of hazard ratios for dementia from joint longitudinal-hazard models for *APOE* e4 alleles, years of education, and child cognitive function. Estimates are standardized so that hazard ratios are expressed for *APOE* per e4 allele, years of education per 4 years of education, and child cognitive function per SD. Error bars represent 95% confidence intervals for hazard ratios. The black dashed line indicates a hazard ratio of 1, where risk for dementia is not affected by the predictor. In both samples, having 1 *APOE* e4 allele significantly increases the risk of dementia via cognitive change, whereas the education reduces the risk of dementia primarily via cognitive levels.

We found consistent results across LBC1936 and EUR-like ancestry participants in HRS (Figure 1). Consistent with meta-analyses and reviews implicating educational attainment in baseline cognitive function, but not rate of longitudinal cognitive decline (Lövdén et al., 2020; Seblova et al., 2020), educational attainment was strongly associated with lower dementia hazard via baseline adult cognitive function in both samples (HR_HRS_ = .43 [.40-.45], HR_LBC_=.44 [.27-.72]) but negligibly associated with dementia hazard via longitudinal slope (HR_HRS_ = .90 [.83-.98], HR_LBC_=1.22 [.89-1.68]). Conversely, consistent with a very large body of empirical research (e.g. Caselli et al., 2009; Deary et al., 2002), *APOE* e4 allele count was strongly associated with dementia hazard via longitudinal cognitive slope (HR_HRS_ = 1.88 [1.59-2.23], HR_LBC_=1.73, [1.31-2.15]) but not baseline cognitive function (HR_HRS_ = 0.99 [.92-1.06], HR_LBC_=1.05, [.99-1.12]). Moreover, as expected, childhood cognitive function (only available in LBC1936) was related to lower dementia hazard exclusively via baseline adult cognitive function (HR_LBC_=.75 [.61-.91]). The consistency of estimates across both samples, and the confirmation in LBC1936 that childhood cognitive function effects operate via baseline adult cognitive function rather than aging-related cognitive change, supports the implementation of joint longitudinal-hazard modeling to make inferences with respect to the life course timing of a broader set of dementia risk factors in the HRS.

To illustrate how these distinct patterns of associations with baseline cognitive function and longitudinal cognitive change can both give rise to AD/ADRD onset, we plot the distributions of HRS model-implied trajectories stratified by different levels of educational attainment and *APOE* genotype in Figure 2. When stratifying by effects of educational attainment on baseline levels, considerable heterogeneity is observed, such that participants with lower educational attainment, on average, reach MCI and dementia thresholds earlier than those with higher levels of educational attainment. However, when stratifying by effects of educational attainment on rates of longitudinal change, differences in educational attainment are not differentiated, and no appreciable difference in rate of dementia hazard is observed. The converse pattern is observed for *APOE*: differences in e4 allele count are associated with rates of dementia by way of differences in longitudinal rates of cognitive change but not differences in baseline levels of cognitive function.

**Figure 2.**
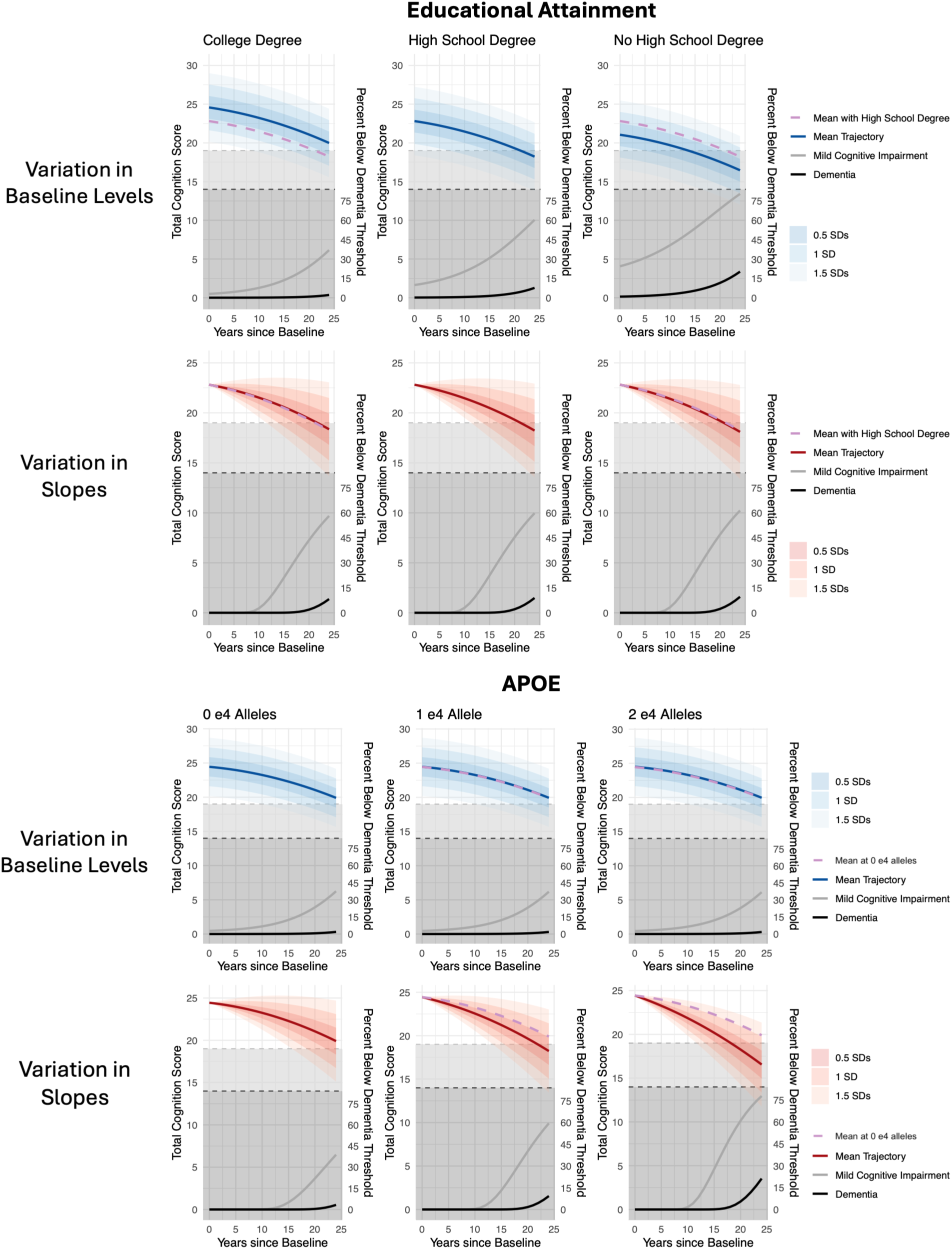
Longitudinal trajectories over time in decades for total cognition scores showing the effect of education and *APOE* on the levels (blue) and slopes (red) of cognitive scores in the Health and Retirement Study. All panels show change in total cognition scores over time. The threshold for a designation of mild cognitive impairment (MCI) is represented by the gray dashed line, and the threshold for a designation of dementia is represented by the black dashed line. The shaded regions around the trajectories represents the distribution of variation in either baseline levels or rates of change within each group. The lower region of each panel represents the proportion of individuals meeting criteria for designation of MCI and dementia, at each age. The trajectories in blue stratify baseline levels of cognitive function by level of the covariate, holding constant longitudinal rates of change. The trajectories in red stratify longitudinal rates of cognitive change by level of the covariate, holding constant baseline levels When stratifying by effects of educational attainment on baseline levels, considerable heterogeneity is observed, such that participants with lower educational attainment, on average, reach MCI and dementia thresholds earlier than those with higher levels of educational attainment. However, when stratifying by effects of educational attainment on rates of longitudinal change, differences in educational attainment are not differentiated, and no appreciable difference in rate of dementia hazard is observed. The converse pattern is observed for *APOE*: differences in e4 allele count are associated with rates of dementia by way of differences in longitudinal rates of cognitive change, but not differences in baseline levels of cognitive function.

### Genetic Predictors of Dementia in HRS

We assessed APOE e4 status and multi-ancestry polygenic indices (PGIs) in three major ancestry groups available in HRS; European-like ancestry (EUR, N=8,728), African-like ancestry (AFR, N=1,608), and Admixed American ancestry (AMR, N=1,182). We identified large scale GWAS of risk factors corresponding to the *Lancet* commission report, which we used to construct polygenic indices (Modifiable risk factors outlined in Table S9, GWAS described in Table S10 and **Online Methods**). We also included PGIs for general cognitive function, AD and for two additional phenotypes not included in the *Lancet* report, but for which very highly powered GWAS are available, and have been implicated in risk for AD/ADRD: externalizing (N_EUR_=1,492,085), which describes a broad dimension of health-relevant risk-taking behaviors; and schizophrenia (N_EUR_=67,390ca/94,015ctrl), polygenic load for which has been linked with cognitive aging in unaffected individuals. Source of GWAS are listed in the **Online Methods**. These PGIs have several desirable properties that position them to provide complementary information to the measured epidemiological risk factors: a) they can be used to proxy risk factors that were not directly measured or only measured using self-report in the prediction sample, b) they index latent risk that may not be fully realized in a measurable risk factor, and c) they do not change during disease progression (and only index genetic effects on pre-morbid factors when trained using GWAS of cognitively healthy individuals). Nevertheless, it is important to note that genetics only account for a portion of total variation in each exposure, and PGIs only index a portion of that genetic variation. Thus, the effect sizes for PGI analyses are expected to be considerably lower than those for direct epidemiological measures.

Results for longitudinal-hazard modeling with genetic predictors are shown in Figure 3 and complete parameter estimates are presented in Supplementary Tables S2-S4. Although APOE e4 substantially increases dementia hazard via cognitive change in EUR participants (HR_s-EUR_ = 1.88 [1.59-2.23]), it was not significantly associated with dementia hazard via premorbid differences or cognitive change in AFR and AMR groups. Hazard ratios for APOE e4 via cognitive change were substantially smaller in the AFR and AMR groups, indicating that it may be of greater relevance for dementia risk in EUR individuals. (HR_s-AFR_ = 1.16 [.95–1.43], HR_s-AMR_=1.14 [.92-1.42]). The Alzheimer’s PGI (excluding the APOE region) was associated with dementia hazard via steeper rates of decline in the EUR and AMR groups, highlighting that the genetics of Alzheimer’s disease index primarily degenerative processes (HR_i-EUR_ = 0.98 [.95– 1.01], HR_s-EUR_=1.21 [1.14-1.27], HR_i-AMR_ = 1.03 [.92–1.15], HR_s-AMR_=1.15 [1.01-1.32]). The Alzheimer’s disease PGI was not significantly associated with dementia hazard through either pathway in the AFR group (HR_i-AFR_ = 1.05 [.95–1.15], HR_s-AFR_=1.03 [.90-1.18]). Also of note is that the general cognitive function PGI was largely associated with reduced dementia hazard via higher baseline levels in EUR, AFR, and AMR groups (Figure 3, Tables S3-S5). PGIs for educational attainment were also significantly associated with reduced dementia hazard via premorbid cognitive differences in EUR and AFR participants (HR_i-EUR_ = .74 [.71–.76], HR_i-AFR_ = .83 [.75–.92]), but this association was attenuated and consequently nonsignificant in the AMR group (HR_i-AMR_=.92 [.83-1.03]).

**Figure 3.**
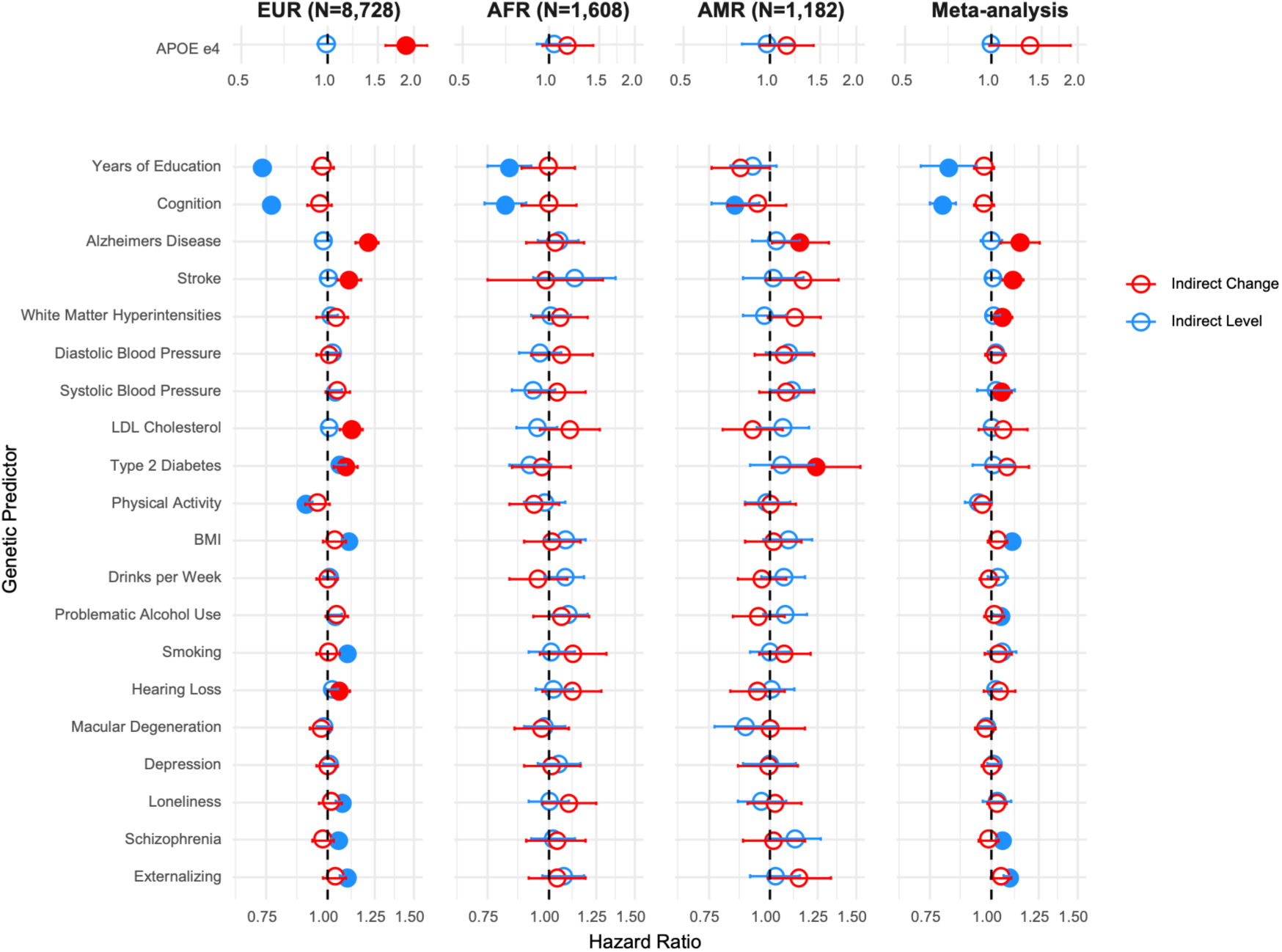
Dementia hazard ratios for genetic predictors via cognitive change (red) and baseline level (blue). Estimates of hazard ratios for dementia from joint longitudinal-hazard models for genetic predictors in EUR, AFR, and AMR samples. Meta-analytic hazard ratio estimates across all 3 groups are shown in the far right panel. Meta-analytic estimates were calculated using an inverse-variance weighted random effects model. For all predictors except *APOE*, which is expressed per e4 allele, hazard ratios are expressed in per SD units. Error bars represent 95% confidence intervals for hazard ratios. Non-significant associations (p > 0.05) are shown in white with outlines corresponding to their pathway.

The majority of remaining PGIs in AFR and AMR ancestry groups were not significantly associated with dementia hazard via premorbid differences or aging-related decline but had effect sizes comparable to the EUR group and greater standard errors, consistent with reduced predictive power of PGIs in non-EUR groups. In the EUR sample, polygenic indices for stroke, type 2 diabetes, LDL cholesterol, and hearing loss were associated with dementia hazard via rates of longitudinal cognitive decline. Each PGI was associated with an approximately 6-12% increase in dementia hazard via steeper cognitive decline in the EUR group (Table S2). The PGI for type 2 diabetes was associated with a 6% increase in dementia hazard via lower baseline cognitive function and an additional 9% increase in dementia hazard via steeper cognitive decline in the EUR group (HR_i-EUR_ = 1.06 [1.03–1.09], HR_s-EUR_=1.09 [1.03-1.15]), a 24% increase in dementia hazard via steeper cognitive decline in the AMR group (HR_s-AMR_=1.24 [1.01-1.53]; Table S4), and not significantly associated with dementia hazard in the AFR group. Additionally, the hearing loss PGI was significantly associated with increased dementia risk only via steeper rates of cognitive decline in EUR participants. In cross-ancestry meta-analysis, PGIs for AD, stroke, white matter hyperintensities, and systolic blood pressure were significantly associated with dementia hazard via rates of cognitive change, and PGIs for BMI, problematic alcohol use, schizophrenia, and externalizing were related to dementia hazard via baseline levels of cognitive function. PGIs for diastolic blood pressure, macular degeneration, drinks per week, and depression were not significantly associated with dementia hazard via either baseline level or longitudinal change, although we caution against concluding that polygenic propensity for these factors is unrelated to dementia hazard. PGIs based on larger discovery samples will be more highly powered to detect such associations.

### Epidemiological Predictors of Dementia in HRS

The *Lancet* 2024 Commission report identified 14 modifiable risk factors for dementia risk: education, hearing loss, high LDL cholesterol, depression, traumatic brain injury, physical inactivity, diabetes, smoking, hypertension, obesity, excessive alcohol consumption, visual loss and air pollution. The HRS contains measures of nearly all these modifiable risk factors (Supplementary Table S9) except traumatic brain injury. Measures of air pollution are not publicly available in the HRS so we were unable to include them in these analyses. Importantly, we use measurements at baseline for epidemiological risk factors, where participants are age 51 or older (M_age_ = 68), although risk factors may be most modifiable earlier in the life course. We examined whether the measure of each modifiable risk factor was associated with dementia hazard via premorbid cognitive differences and differences in cognitive change. Results for epidemiological factors are shown in Figure 4 and complete parameter estimates are reported in Supplementary Table S5-S8. We focus here on results for the full sample (right panel of Figure 4) and report results separately by demographic group (European-like ancestry sample, self-reported black sample, self-reported Hispanic sample) to evaluate the consistency of estimates across these groups (first, second, and third panels of Figure 4). It is of note that, because subgroup analyses are necessarily lower powered, it is most sensible to compare overall patterns of effects, as opposed to comparing the consistency of significance levels, across subgroups. Patterns were generally consistent across these subgroups, but we point out some notable differences.

**Figure 4.**
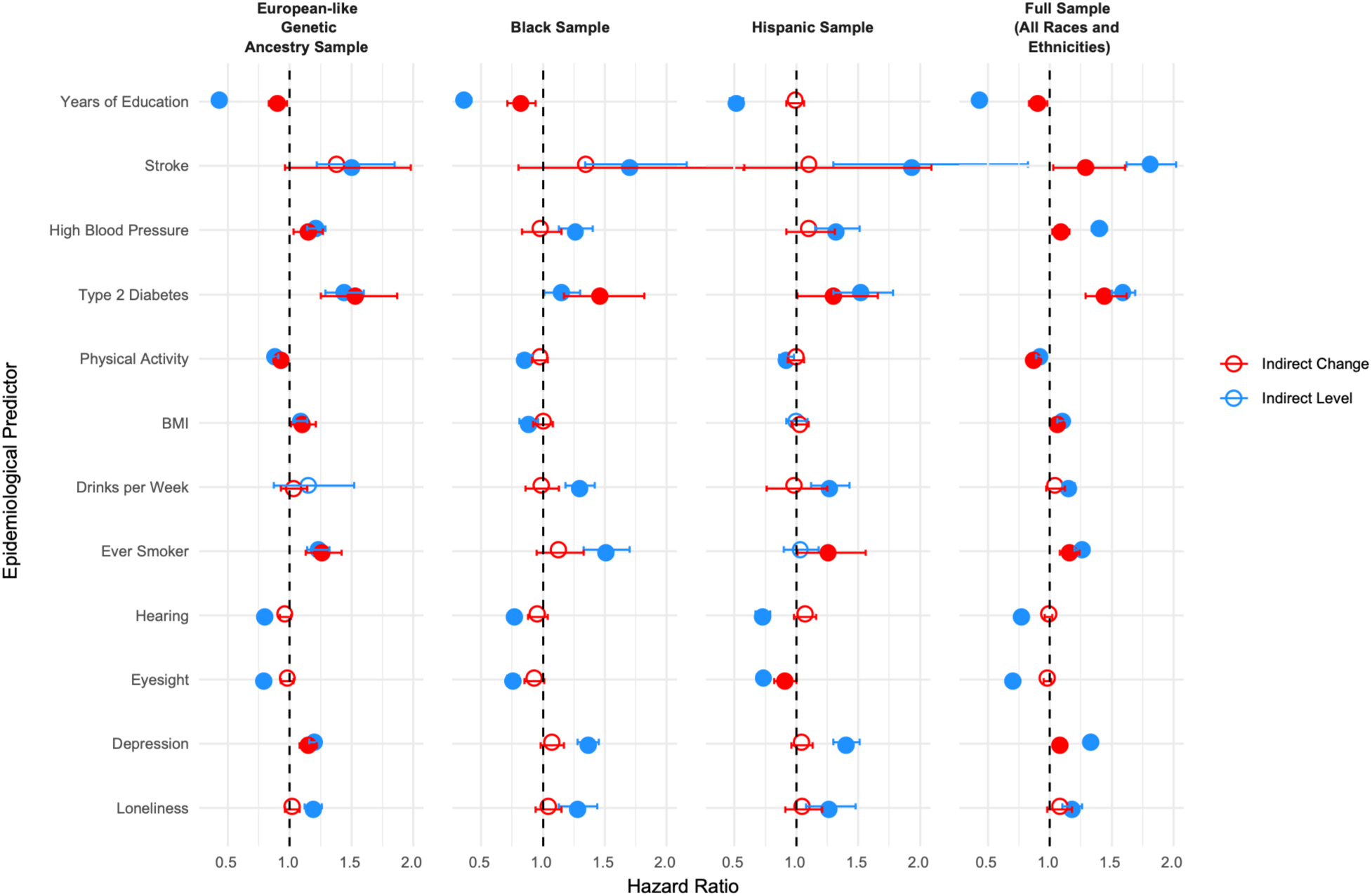
Dementia hazard ratios from joint longitudinal-hazard models for epidemiological predictors via cognitive change (red) and baseline level (blue) in the full sample and specific demographic groups. For all predictors except years of education hazard ratios are expressed in per SD units. Hazard ratios for years of education are expressed per 4 years of education. Error bars represent 95% confidence intervals for hazard ratios. Non-significant associations (p > 0.05) are shown in white with outlines corresponding to their pathway.

All epidemiological risk factors were prospectively related to dementia hazard, yet displayed considerable heterogeneity in the extent to which effects were mediated by baseline levels or longitudinal rates of cognitive change. Beyond educational attainment and *APOE* genotype, Type 2 diabetes was among the strongest predictors of dementia hazard, with effects occurring similarly across both baseline levels and rate of change. A Type 2 diabetes diagnosis was associated with an approximately 50% increase in dementia hazard via lower baseline cognitive function and an additional 50% increase in dementia hazard via steeper rate of cognitive decline (HR_i_ = 1.59 [1.5 – 1.69], HR_s_=1.44 [1.29-1.62]). Higher BMI, less physical activity, and smoking were also associated with dementia hazard at similar levels for baseline levels and rate of change (Table S5). One SD higher BMI was associated with an approximately 10% increase in dementia risk via lower baseline cognitive function and an additional 6% increase in dementia hazard via steeper rate of cognitive decline (HR_i_ = 1.10 [1.06 – 1.14], HR_s_=1.06 [1.01-1.11]). In the Black subsample higher BMI was protective against dementia, with effects occurring via baseline levels, possibly reflecting a survivor bias or what has sometimes been referred to as the obesity paradox of aging (e.g., Childers & Allison, 2010, Table S7). Self-reported smoking history at baseline was associated with a 25% increase in dementia hazard via lower baseline cognitive function, and an additional 15% increase in dementia hazard via steeper cognitive decline (HR_i_ = 1.26 [1.2 – 1.32], HR_s_=1.16 [1.08-1.24]). One SD higher levels of physical activity was associated with an approximately 15% reduction in risk via higher baseline cognitive function and an additional 5% reduction in risk via shallower rates of cognitive decline.

Higher blood pressure, depression, and stroke occurrence prior to baseline assessment were primarily associated with dementia hazard via lower baseline cognition, with some additional mediation by steeper rates of change. Occurrence of stroke prior to baseline assessment was associated with an 80% increase in dementia hazard via lower baseline cognitive function, and an additional 30% increase in dementia hazard via steeper cognitive decline (HR_i_ = 1.81 [1.62 – 2.02], HR_s_=1.29 [1.03-1.61]). Diagnosis of high blood pressure at baseline was associated with a 40% increase in dementia hazard via lower baseline cognitive function, and an additional 10% increase in dementia hazard via steeper cognitive decline (HR_i_ = 1.4 [1.34 – 1.46], HR_s_=1.09 [1.02-1.16]). Reporting one SD higher depression symptoms was associated with an approximately 30% increase in dementia hazard via lower baseline cognitive function, and an additional 8% increase in risk via steeper cognitive decline (HR_i_ = 1.33 [1.3 – 1.36], HR_s_=1.08 [1.05-1.12]).

Finally, greater alcohol consumption, higher loneliness, and poorer self-ratings of hearing and eyesight were generally only associated with dementia hazard via lower premorbid cognitive function, although there was some evidence that poorer self-ratings of eyesight were also associated with dementia hazard via steeper rate of cognitive decline in the Hispanic subsample. Although the PGI for hearing loss was associated with dementia hazard via steeper rate of cognitive decline in the EUR subsample, phenotypic poorer self-rating hearing was associated with dementia hazard via lower premorbid cognitive function across all groups. In the full sample, one SD greater self-rating of hearing and eyesight were each associated with an approximately 25% reduction in dementia hazard via baseline levels of cognitive function. One higher SD self-reported drinks per week and one SD higher levels of self-reported loneliness were associated with a 15% and 25% increase in dementia hazard via lower baseline cognitive function, respectively.

## Discussion

Clinically impaired levels of cognitive functioning characteristic of AD/ADRD can arise from a combination of steep rates of cognitive decline to a diagnostic threshold of low cognitive function and lower premorbid levels of cognitive functioning that place individuals closer to the diagnostic threshold prior to the aging process. Here, we have capitalized on the joint analysis of longitudinal and time-to-event data to discern between these time-ordered pathways for a range of genetic and epidemiological predictors of AD/ADRD hazard. We found that the APOE e4 allele and PGIs for AD, type 2 diabetes, stroke, LDL cholesterol, and hearing loss were associated with dementia hazard via rates of cognitive change, and PGIs for educational attainment, cognitive function, physical activity, BMI, smoking, loneliness, schizophrenia, and externalizing were associated with dementia hazard via premorbid differences in cognitive function. History of stroke, Type 2 diabetes, BMI, smoking, and physical activity exhibited associations with dementia hazard via both baseline level and longitudinal change. Both depression and high blood pressure were also associated with dementia hazard via longitudinal change, although their associations via baseline levels were appreciably stronger. Hearing, eyesight, loneliness, and alcohol intake were associated with dementia exclusively via baseline levels. These findings have considerable implications for understanding the mechanisms of AD/ADRD risk and determine the most promising periods of the lifespan for application of interventions targeting each of these factors.

Notably, effects varied by ancestry group. For example, PGIs for AD and Type 2 Diabetes were similarly associated with dementia hazard via cognitive change in the EUR and AMR groups, but were not significantly associated with dementia hazard via cognitive change in the AFR group. Given the relatively lower samples sizes of the AFR GWAS contributing to the multi-ancestry PGIs, lower PGI prediction may be at least partially explained by issues of portability related to differences in minor allele frequency and linkage disequilibrium structure across ancestry groups (Wang et al., 2020). Similarly, some PGIs (e.g. macular degeneration, white matter hyperintensities) were computed based on GWAS with limited sample size in all ancestries, including EUR. This may explain why PGIs for some risk factors had null or small associations with to dementia hazard.

While we validated our analyses by demonstrating very consistent results for educational attainment and *APOE* in HRS and the deeply phenotyped LBC1936, the sample size of LBC1936 did not permit a highly powered examination of the broader range of risk factors that we examined in the HRS. Langa-weir scores have been extensively validated and used in other research (Crimmins et al., 2011; Langa et al., 2005, 2017; Wei et al., 2018), but may have key differences from clinically diagnosed AD/ADRD. Future work in large deeply phenotyped cohorts or consortia may provide more mechanistic insight into the links between modifiable risk factors and cognitive aging processes over time.

An association between a baseline risk factor and dementia hazard via subsequent cognitive change may arise, not because that risk factor causally affects cognitive decline, but because it represents an early indicator of disease progression that forecasts but does not causally affect decline. Lower cognition in early life may itself be a risk factor for subsequent changes in lifestyle, health behaviors, and physical health (Deary et al., 2021; Twig et al., 2018). Although the analyses reported here excluded individuals with dementia at baseline, it is likely that many participants experienced undetected Alzheimer’s pathology-related neurocognitive change for years prior to the baseline assessment. Thus, although we consider the risk factors identified to be promising targets for future research efforts to prevent dementia, their causal status and biological mechanisms of action currently remain ambiguous. Randomized controlled trials are needed to obtain decisive conclusions regarding the effects of these risk factors and their utility as intervention targets to prevent, delay, or treat AD/ADRD.

In conclusion, we implemented joint longitudinal-hazard models to distinguish two distinct pathways via which genetic risk for dementia operates. Factors that operate via differences in cognitive aging may be the most promising targets to investigate in future efforts to develop later-life interventions to slow cognitive decline or prevent clinical impairment.

## Data Availability

Data used in the present study are available by application to the Health and Retirement Study and Lothian Birth Cohort.

## Online Methods

### Participants and Measures

#### The Health and Retirement Study

The Health & Retirement Study (HRS) is a nationally-representative (of the USA) sample of adults over age 50 years (and their spouses, regardless of age) that includes oversamples of Hispanic and Black participants (Sonnega et al., 2014). The sample for these analyses (N=30,509) contains adults that participated in cognitive testing in at least one of 14 HRS timepoints (every two years from 1996-2020) and those that participated in cognitive testing in the AHEAD sample in 1995, yielding up to 25 years of longitudinal cognitive data per person. Participants aged 50 years and younger were excluded. Data from the 1992 and 1994 waves of HRS were excluded because stimuli for some cognitive tasks administered in these waves were not the same as those from later waves. Participants receiving Langa-Weir scores corresponding to MCI or dementia at baseline were excluded. Supplementary Table S11 reports basic descriptive statistics for participants at their first wave of cognitive testing.

##### Cognitive Measures

The HRS cognitive battery includes 7 measures of cognitive function: Immediate and delayed word recall, serial 7s, backwards counting, dates, naming, and vocabulary. Details on administration of each cognitive task can be found in Herzog & Wallace (1997). We used total cognition score (ranges from 0-35) which is the sum of scores on the immediate recall (0-10 points), delayed recall (0-10 points), serial 7s (0 to 5 points), backwards counting (0-2 points), dates (0-4 points), and president and object naming tasks (0-2 points each). HRS participants completed all 7 cognitive tasks at baseline and at follow-up time points at which they were 65 years and older. Additionally, the telephone interview for cognitive status (TICS) score includes serial 7s, backwards counting, date naming, object naming, and president/vice president naming tasks, which are assessed for all participants at all time points, regardless of age. Primary models were fit to the total cognition composite (a continuously distributed score without evidence for ceiling or floor effects; Figure S1). We also conducted a set of sensitivity analyses using immediate recall scores (also normally distributed with minimal evidence of ceiling or floor effects) to test sensitivity of results to modeling changes specifically associated with episodic memory and a set of sensitivity analyses using the TICS score to test the sensitivity of results to the availability of more frequent follow-up measures between ages 51 and 65 (**Supplementary Note**). Cognitive assessment data from time points concurrent with or after a dementia onset were set to missing to specifically estimate how rates of premorbid cognitive change relate to subsequent dementia hazard. We did not include imputed values for the cognitive measures or cognitive data from proxy respondents. As Langa-Weir scores were only available in 1995 and later timepoints, we did not include cognitive scores from 1993. Participants whose original first testing date was in 1992, 1993, or 1994 were assigned a baseline testing date as the earliest cognitive testing they participated in that was included (e.g. 1995 or 1996).

##### Dementia Ascertainment

Dementia is classified in the HRS using Langa-Weir scores, described extensively in previous publications (Crimmins et al., 2011; Langa et al., 2005, 2019). Langa-Weir scores were developed by mapping scores on the HRS cognitive measures to cognitive states ascertained in the Aging, Demographics, and Memory Study (ADAMS) sample by a consensus panel based on 3-4 hour structured home-based interviews by a team consisting of a nurse and neuropsychology technician. When a participant is unable to respond for themselves, Langa-Weir scores are based on information from proxy respondents, who complete a direct assessment of memory and an assessment of limitations in five instrumental activities of daily living and an interviewer assessment of difficulty completing the interview due to cognitive limitation. Langa-Weir classifications range from 1-3, where 1 indicates normal cognitive functioning, 2 indicates cognitive impairment but not dementia, and 3 indicates dementia. Langa-Weir classifications in HRS correspond with the gold standard assessments made in ADAMS, particularly for individuals that are cognitively normal and those diagnosed with dementia, and Langa-Weir scores are routinely used in epidemiological research (e.g. Davydow et al., 2015; Langa et al., 2017; Wei et al., 2018). We classify participants that receive a Langa-Weir score of 3 as meeting criteria for incident dementia, and thus code all future Langa-Weir scores for that participant as 3. For hazard analyses, participants are considered right-censored if they dropped out of the study prior to receiving a Langa-Weir score of 3. Across all years of measurement, 30-40% of all dementia cases in that year were based on proxy data, and 30-40% of initial instances of dementia classification were based on proxy data (Figure S2, Figure S3). Note that we also perform a validity analysis that compares results for *APOE* e4 status and educational attainment (two canonical predictors of dementia hazard) to those from the LBC1936 in which AD/ADRD is ascertained by a team of clinical dementia experts, and cognitive function is measured using an extensive in-person neurocognitive test battery.

### Confirmatory Predictors

We selected two well-established predictors of dementia risk to validate the longitudinal-hazard approach: Educational attainment and number of *APOE* e4 alleles. Education was coded as number of completed years of schooling (Supplemental Table S12), to account for differences between school systems in the United States and Scotland (Miller, 2007), and divided by four such that one unit corresponds to a 4-year degree. The e4 allele of the *APOE* gene is well-known risk factor for AD/ADRD that occurs commonly within the general population (Corder et al., 1993; Serrano-Pozo et al., 2021; Strittmatter & Roses, 1996; William Rebeck et al., 1993). *APOE* status was coded where 0 indicated the participant had no e4 alleles, 1 indicated the participant had one e4 allele and 2 indicated the participant had 2 e4 alleles. Of the 16,802 participants with *APOE* data, 4288 (26%) carried 1 e4 allele and 373 (2%) carried 2 e4 alleles. The majority of participants had APOE directly genotyped, with APOE genotyped imputed from SNP array data for small proportions of participants. APOE was imputed for 1,090/8,796 of EUR participants, 225/2,329 of AFR participants, and 132/1,766 of AMR participants.

### Polygenic Predictors

Polygenic indices (PGIs; Becker et al., 2021) are individual summary scores of weighted associations between genetic variants and a given outcome, weighted by effect size estimates derived from a large external Genome Wide Association Study (GWAS) that did not include the prediction sample (here, the HRS; we use summary statistics that exclude the HRS). We selected the most recent EUR GWAS corresponding to the modifiable risk factors identified in the 2024 Lancet Commission on Dementia report (Livingston et al., 2024) in addition to other factors that have been separately implicated in dementia risk (i.e. externalizing, schizophrenia). We selected corresponding AFR and AMR GWAS with sample sizes greater than 3,000 or, for traits where EUR N < 100,000, we use GWAS data where AFR or AMR sample size is greater than 5% of the EUR sample size. We use the following GWAS: educational attainment (N_EUR_=3,026,656; Okbay et al., 2022), general cognition (N_EUR_= 274,617; Davies et al., 2018), hearing loss (N_EUR_=147,997ca/575,269ctrl; Trpchevska et al., 2022); LDL cholesterol (N_EUR_=1,097,570, N_AFR_=262,383, N_AMR_=58,398; Graham et al., 2021); depression (N_EUR_=658,808ca/4,364,225ctrl; Adams et al., 2025, N_AFR_=9.649ca/122,347ctrl, N_AMR_=22,927ca/340,403ctrl; Meng et al., 2024); stroke (N_EUR_=110,182ca/1.5Mctrl, N_AFR_=3,691ca/20,030ctrl, N_AMR_=1,516ca/4,146ctrl; Mishra et al., 2022), white matter hyperintensities (N_EUR_=50,970; Sargurupremraj et al., 2020); moderate-to-vigorous physical activity (N_EUR_=608,595; Wang et al., 2022); type 2 diabetes (N_EUR_=428,452ca/2.1Mctrl, N_AFR_=50,251ca/103,909ctrl, N_AMR_=29,375ca/59,368ctrl; Suzuki et al., 2024); smoking (N_EUR_=557,337ca/674,754ctrl, N_AFR_=8,571ca/13,427ctrl; Saunders et al., 2022); systolic and diastolic blood pressure (N_EUR_=1,028,980; Keaton et al., 2024, N_AFR_=119,274, N_AMR_=57,988; Verma et al., 2024), BMI (N_EUR_=681,275; Yengo et al., 2018, N_AFR_=118,993, N_AMR_=57,793; Verma et al., 2024); problematic alcohol use (N_EUR_=903,147, N_AFR_=40,116ca/82,455ctrl; Zhou et al., 2023), alcohol consumption (N=2,428,851, N_AFR_=7,072, N_AMR_=5,162; Saunders et al., 2022); loneliness (N_EUR_=456,422; Jiang et al., 2019); early age related macular degeneration (N_EUR_=14,034ca/91,214ctrl; Winkler et al., 2020, N_AFR_=7,126ca/109,352ctrl, N_AMR_=4,642ca/52,515ctrl; Verma et al., 2024); Alzheimer’s Disease (excluding the *APOE* region (Chr19:45,000,000-45,800,000, GRCh37/hg19), as *APOE* genotype is entered as a separate predictor (N_EUR_=100,806ca/2,026,274ctrl, N_AFR_=4,379ca/66,731ctrl; Uffelmann et al., 2026). We also created PGIs for two additional behavioral/psychiatric phenotypes not included in the Lancet report, but for which very highly powered GWAS are available, and have been implicated in risk for AD/ADRD (Cai & Huang, 2018; Ritchie et al., 2020; Solares et al., 2023): externalizing (N_EUR_=1,492,085; Karlsson Linnér et al., 2021), which describes a broad dimension of health-relevant risk-taking behaviors; and schizophrenia (N_EUR_=67,390ca/94,015ctrl, N_AFR_=6,152ca/3,918ctrl; Trubetskoy et al., 2022), polygenic load for which (even among unaffected individuals) has been linked with cognitive aging.

Although GWAS data for AFR and AMR groups fitting our criteria were available for hearing loss (Verma et al., 2024), these data were primarily from the Million Veteran Program (MVP), a cohort of military members, which may be especially more likely to experience hearing loss due to non-aging related factors. The EUR GWAS of hearing loss from Verma et al., 2024 and the Trpchevska et al., (2022) GWAS of hearing loss were genetically correlated at 0.63, suggesting that there may be meaningful differences in the etiology of hearing loss between these two cohorts. As a result, we elected to only use the EUR GWAS data from Trpchevska et al., (2022) and did not compute multi-ancestry PGIs for hearing loss.

Genotype array data necessary for computing PGIs are available for n=8,728 non-Hispanic White participants, n=1,608 AFR participants and n=1,182 AMR participants. Primary global ancestry was determined in HRS using self-reported race and ethnicity in addition to individual principal component loadings from principal component analysis of all study subjects. The EUR sample includes self-reported non-Hispanic whites with PC loadings within 1 standard deviation of the mean for eigenvectors 1 and 2 in PC analysis. The AFR sample includes self-reported African Americans within 2 standard deviations of the mean of all self-identified African Americans for eigenvector 1 and 1 standard deviation of the mean for eigenvector 2 in PC analysis of all subjects. The AMR sample includes all self-reported Hispanic participants with outliers removed that fell outside 6 standard deviations of the mean for PCs 1-10.

For prediction in EUR samples, or in cases where only EUR GWAS data was available, we use SBayesRC to derive PGI weights. SBayesRC (Zheng et al., 2024) is a Bayesian method that incorporates functional annotation data to distinguish likely causal SNPs from non-causal SNPs. SBayesRC uses a low-rank model to model the LD among common variants, then uses a multicomponent annotation-dependent mixture prior to model the distribution of SNP effect sizes. For each trait, we estimated PGI weights with SNPs sorted into five using probability starting values of 95%, 2%, 2%, and 1%, with effect sizes scaled at .00, .01, .10, and 1.00. We used BaselineLD v2.2 genomic annotation data (Gazal et al., 2017) and approximately 1 million common SNPs from HapMap3 for LD reference (Zheng et al., 2024). The MHC region was excluded from these analyses. We used the SBayesRC weights to estimate a score for each participant using PLINK2 (Chang et al., 2015).

PGIs computed from European-like ancestry (EUR) summary statistics typically produce attenuated predictive performance in other ancestries. However, GWAS data in non-EUR ancestries is often limited (Privé et al., 2022; J. Y. Wang et al., 2026). To maximize prediction in AFR and AMR samples where AFR and AMR GWAS data were available (according to specifications outlined above), we generated multi-ancestry PGI weights using PRS-CSx, a Bayesian method which couples genetic effects across populations by estimating a shared continuous shrinkage prior (Ruan et al., 2022). We used PRS-CSx auto to derive multi-ancestry PGI weights integrating data from the EUR GWAS and relevant ancestry group (AFR or AMR) corresponding to each non-EUR participant’s primary global ancestry. PRS-CSx auto derives PGI weights from continuous shrinkage priors shared across the two ancestry groups (i.e. EUR and AFR or EUR and AMR). The “auto” option develops global shrinkage priors directly from GWAS summary statistics, bypassing the need for a tuning sample. We used the “meta” option, where PRS-CSx combines SNP posterior effect sizes from both ancestry groups using an inverse-variance-weighted meta-analysis to derive trans-ancestry PGI weights. We used the PRS-CSx weights to estimate PGIs for each HRS participant using PLINK2 (Chang et al., 2015).

In all models with genetic factors, we included principal components of ancestry to avoid confounds associated with population stratification. PGIs were standardized prior to analysis.

### Epidemiological Predictors of Dementia Hazard

We selected epidemiological predictors available in the HRS that correspond with the modifiable risk factors identified in the most recent Lancet Dementia Commission Report (Livingston et al., 2024), which include education, hearing loss, high LDL cholesterol, depression, physical inactivity, diabetes, smoking, hypertension, obesity, excessive alcohol use, social isolation, visual loss, and air pollution. We use the measurement of each predictor concurrent with or prior to the baseline cognitive assessment to afford prospective prediction of change and dementia hazard. For measures introduced to the HRS at later waves, we removed cognitive data from waves preceding that measurement (Years each epidemiological predictor was assessed are noted in Table S9). Hearing and Vision are self-rated (from 1 = “poor” to 5 = “excellent”). Depression is measured by the 8-item CESD (Karim et al., 2015). Physical activity is measured using self-report of frequency of engagement in vigorous physical activity (1 = “hardly ever or never” to 5 = “every day”). Alcohol consumption is calculated as the number of days per week a participant reports drinking alcohol multiplied by the average number of drinks consumed per day. Social Isolation is measured using the sum of 3 items from the UCLA Loneliness scale (Russell, 1996) that assesses the degree to which participants lack companionship, feel isolated from others, and feel left out. Stroke (implicated in the Lancet report, but not included among the list of 14 modifiable risk factors), hypertension, smoking, and type 2 diabetes are measured by participant self-report. Obesity is derived from BMI, based on self-reported height and weight. We were unable to assess air pollution in the HRS as these data were not publicly available.

#### Validation Sample: The Lothian Birth Cohort of 1936

##### Participants

The Lothian Birth Cohort of 1936 (LBC1936) is a narrow-age longitudinal study of aging of community-dwelling adults born in 1936 living in Edinburgh and the Lothians area of Scotland (Deary et al., 2012). All LBC1936 participants are of European-like ancestry. The present study comprises a total of 989 participants from the first five assessments (wave 1, at a mean age about 70 years, to wave 5, at a mean age about 82 years) spanning a total of 12.54 years (Taylor et al., 2018). Of the 1,091 LBC1936 participants measured at baseline, we restricted analyses to participants that had known dementia status and age data for at least one assessment wave, death, or dementia diagnosis, resulting in N=989 participants.

##### Cognitive Measures

LBC1936 participants underwent a battery of 13 cognitive tests representing visuospatial ability, processing speed, verbal memory, and crystallized knowledge (Deary et al., 2012). Cognitive test scores were standardized by subtracting the mean of all data points at the baseline occasion and dividing by the standard deviation of all the data points at baseline. Because we were specifically interested in estimating how rates of premorbid cognitive change related to subsequent dementia onset, 33 data points from cognitive assessments administered concurrent with or subsequent to a dementia diagnosis were set to missing.

##### Dementia Ascertainment

As detailed in Mullin et al. (2023), incident dementia was ascertained based on consensus review board meeting by a multidisciplinary team of clinical dementia experts, drawing on data from electronic health records, death certificates, and home visit clinician assessments. A total of 145 participants in the LBC1936 study received a diagnosis of dementia (15% of the sample), with 46.21% of diagnosed participants being female. Medical records and death certificates were obtained approximately concurrent with wave 5. Thus, for participants who were not classified as meeting criteria for dementia and participated in wave 5, their censoring age is their individual age at wave 5. For participants who were not classified as meeting criteria for dementia and survived until wave 5, but did not participant in wave 5, their censoring age was set as the mean age at wave 5 (82 years). For individuals who were deceased prior to wave 5, but who were not classified as meeting criteria for dementia, we used age at death as the age at censoring. The absolute frequencies of dementia-free participants, dementia-positive participants, and censored cases across age groups are presented in Supplementary Figure S4.

##### APOE status

*APOE*-e4 alleles are a key risk factor for dementia and cognitive decline in older age (Belloy et al., 2019; Blacker et al., 1997; Petersen et al., 1995; Strittmatter & Roses, 1996). Participants had blood samples taken for DNA extraction. *APOE*-e4 status was determined by genotyping the single nucleotide polymorphisms (SNPs), rs429358 and rs7412 using TaqMan technology at the Wellcome Trust Clinical Research Facility Genetics Core at the Western General Hospital in Edinburgh (Luciano et al., 2009). The genotyping was used to determine the presence or absence of *APOE*-e4 alleles. Out of the 934 with available genotyping data, 262 were carriers of one copy of the *APOE*-e4 allele (28%) and 19 were carriers of two copies of the *APOE*-e4 allele (2%).

##### Years of education

Education was coded as number of completed years of formal full-time schooling, to account for differences between school systems in the United States and Scotland (Miller, 2007), and divided by four such that one unit corresponds to a 4-year degree.

##### Child cognitive function

Almost all LBC1936 participants underwent the Scottish Mental Survey of 1947 at approximately 11 years of age, which included the Moray House Test No. 12, a measure of general cognitive function (Deary et al., 2000, 2004). We expected that early life cognitive function would be related to dementia hazard via the growth curve intercept (baseline level of adult cognitive function) and not the slope. The availability of child cognitive function allowed us to explicitly test this key prediction.

#### Joint Longitudinal-Hazard Models

We modelled trajectories of change in cognitive function in relation to dementia hazard using joint longitudinal-hazard models (Ghisletta et al., 2006; Henderson et al., 2000; McArdle et al., 2005). The joint longitudinal-hazard model integrates a growth curve model of variation in baseline level and subsequent rate of longitudinal cognitive change with a hazard model of dementia hazard (**Figure S5)**. By incorporating a time-invariant predictor (measured at or prior to baseline) into the model, the joint model allows us to estimate the extent to which a predictor’s association with dementia hazard acts via effects on baseline cognitive level and longitudinal rate of cognitive change. The growth curve portion of the model can be expressed as follows:

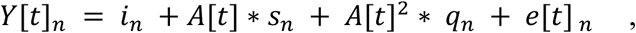

where *Y*[*t*]*_n_* is score *Y* of person *n* on the cognitive measure at time *t*, *i_n_* is the baseline level, *s_n_* is the linear slope, and *q_n_* is the quadratic slope, and *e*[*t*]*_n_* is the time-specific residual for person *n*. The parameter set *A*[*t*] constitutes fixed basis coefficients representing time (in decades) since baseline measurement at each assessment. In LBC1936, the *A*[*t*] coefficients were 0, .298, .675, .0981, and 1.254 for waves 1 through 5, respectively. In HRS, the *A*[*t*] coefficients were specified in constant increments of .2 units, beginning at 0, to reflect the approximate 2-year span between waves. Because the shorter time span of the LBC1936 study does not allow for accurate estimation of curvilinear effects, *q_w,n_* was not estimated in LBC1936. In HRS, the variance of the quadratic slope was fixed to 0 to facilitate model estimation and interpretation. To ensure that the slope terms only capture change preceding dementia onset, all cognitive scores for a given individual concurrent with and after dementia onset were set to missing. In LBC1936, for which multiple high quality cognitive tests were available spanning a total of 4 domains of cognitive function, we fit a multivariate model to estimate factors representing general cognitive function and general change in cognitive function (McArdle, 2009; Ritchie et al., 2016). Additional details on the factor of curves model used in LBC1936 are provided in the supplement.

For the hazard portion of the model, we applied a semiparametric proportional hazard model, allowing for right censoring (study dropout prior to event onset). The regression coefficients that relate external predictors to dementia hazard are expressed as log hazard ratios. Growth curve level (*i*), and slope (*s*), along with covariates *X*_3_to *X_k_* are entered as predictors of dementia hazard as follows:

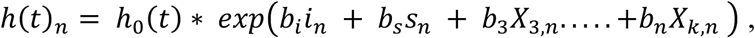

where ℎ_0_(*t*) is the baseline risk of dementia diagnosis at time *t, b_i_* and *b_s_* are the regression coefficients for cognitive level and cognitive linear change (respectively), and *b*_3_*X*_3….._+*b_k_X_k_* refers to additional predictors and covariates (*X_k_*) and their associated regression coefficients (*b_k_*). The slope and intercept terms are also regressed on predictors (e.g. years of education) and additional covariates as follows:

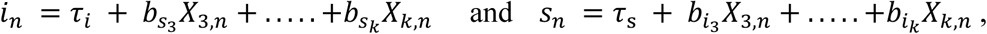

where *τ* terms are regression intercepts and *b* terms are regression coefficients. Models with genetic variables (e.g. *APOE*, PGIs) included the first 10 genetic principal components as covariates to control for population stratification. Age at baseline and year of baseline measurement were included as covariates in all models to account for age and cohort effects. All models control for biological sex. Importantly, the growth curve and hazard portions of the model are simultaneously fit as a single joint model. In other words, we do not output empirical bayes estimates of growth curve slopes for entry into the hazard analysis in a second step, as such an approach can reduce power and introduce bias. Additionally, because levels are considered as simultaneous predictors, their estimated effects on the log dementia hazard are incremental of one another and additive. Analyses were performed using M*plus* software (Muthén & Muthén, 1998–2017), employing full-information maximum likelihood estimation to take all available data into account in the presence of participant dropout.

## Acknowledgements

This research was supported by National Institutes of Health (NIH) grant R01AG073593. MLCS received funding support from NIH grant T32DA007261. EMTD is a member of the Population Research Center (PRC) and the Center on Aging and Population Sciences (CAPS) at The University of Texas at Austin, which are supported by NIH grants P2CHD042849 and P30AG066614, respectively. We thank K.P. Harden (University of Texas at Austin), M. Hayward (University of Texas at Austin), A. Haley (University of Texas at Austin), and Y. Dragostinov (University of Texas at Austin) for their invaluable feedback on earlier versions of this work.

