## Supplementary Note for "Joint Longitudinal Hazard Modeling Distinguishes Polygenic Effects on Dementia via Premorbid Cognitive Function and Aging-Related Decline"

**Supplemental Results**

***Results of Sensitivity Analyses with Different Dementia Coding***

Due to the high proportion of dementia diagnosis based on proxy data, we performed a sensitivity analysis to compare longitudinal-hazard model results for the two confirmatory predictors (Years of Education and APOE genotype) in samples where proxy data was and was not included. Results are shown in Table S13. Models that included the proxy data produced very similar results to models where proxy data was excluded. We therefore elected to include dementia ascertainment scores based on proxy data in our primary analyses

HRS also measures dementia prevalence using self-reported dementia diagnosis. Figure S6 displays the percent of dementia cases by Langa-Weir score or self-report for levels of education and by age. Both dementia measures evince substantially reduced prevalence rates for participants with a high school degree or higher, with further reductions for additional degrees earned. As expected, dementia prevalence also increases with age for both Langa-Weir scores and self-report. Dementia rates are lower for self-reported cases than when measured with Langa-Weir scores.

***Results of Sensitivity Analyses with TICS instead of Total Cognition***

HRS participants completed all 7 cognitive tasks composing the total cognition score at baseline and at follow-up time points at which they were 65 years and older. The telephone interview for cognitive status (TICS) score includes serial 7s, backwards counting, date naming, object naming, and president/vice president naming tasks, which are assessed for all participants at all time points, regardless of age. We conducted sensitivity analyses using the TICS score to test the sensitivity of results for our key confirmatory predictors (Years of Education and APOE genotype) to the availability of more frequent follow-up measures between ages 51 and 65. Parameter estimates are reported in Table S14 and were generally similar between the TICs and total cognition score models.

***Results of Sensitivity Analyses with Immediate Recall instead of Total Cognition***

Parameter estimates from the longitudinal-hazard models estimated using immediate recall scores to measure cognitive levels and slopes are presented in Supplementary Table S15 (epidemiological predictors) and Supplementary Table S16 (genetic predictors). Generally, results were very similar for both phenotypic and polygenic predictors between the immediate recall and total cognition models.

Stroke, BMI, depression and PGIs for stroke and hearing loss, which evinced significant effects on change in total cognition scores, were not significantly associated with change in immediate recall scores. Additionally, in the immediate recall models, drinks per week was significantly associated with increased dementia risk via both cognitive level and cognitive change, whereas in the total cognition model it was only associated with dementia risk via cognitive levels.

Finally, PGIs for alcohol dependence, hearing loss, and schizophrenia all had significant associations with dementia risk via immediate recall levels but did not have significant associations with total cognition levels. These results may indicate that some associations operate more specifically on individual domains of cognitive function.

**Supplementary Figures**

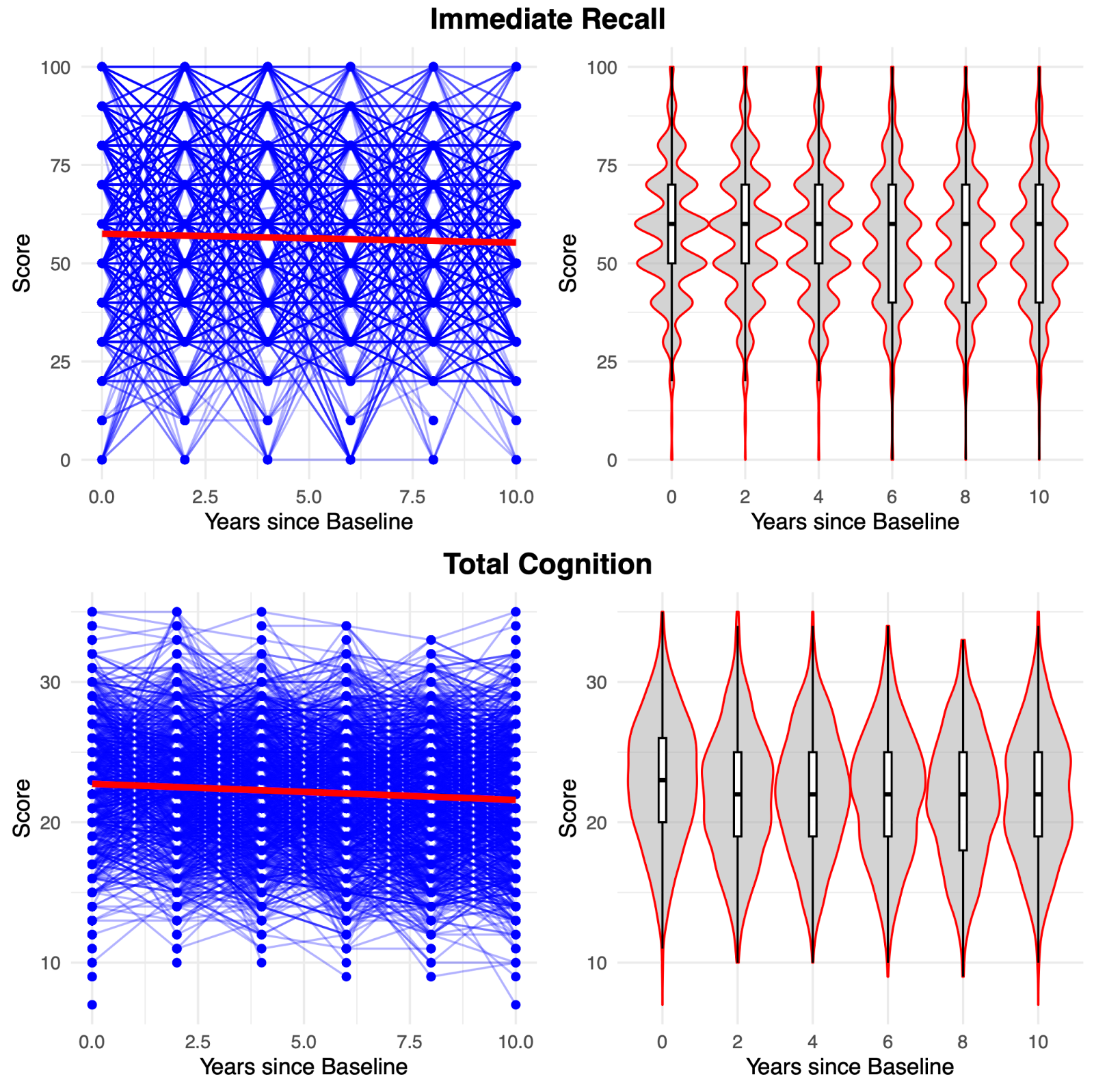

**Figure S1.** Distribution of immediate recall and total cognition scores for a random subsample of 5000 participants across assessment waves in HRS

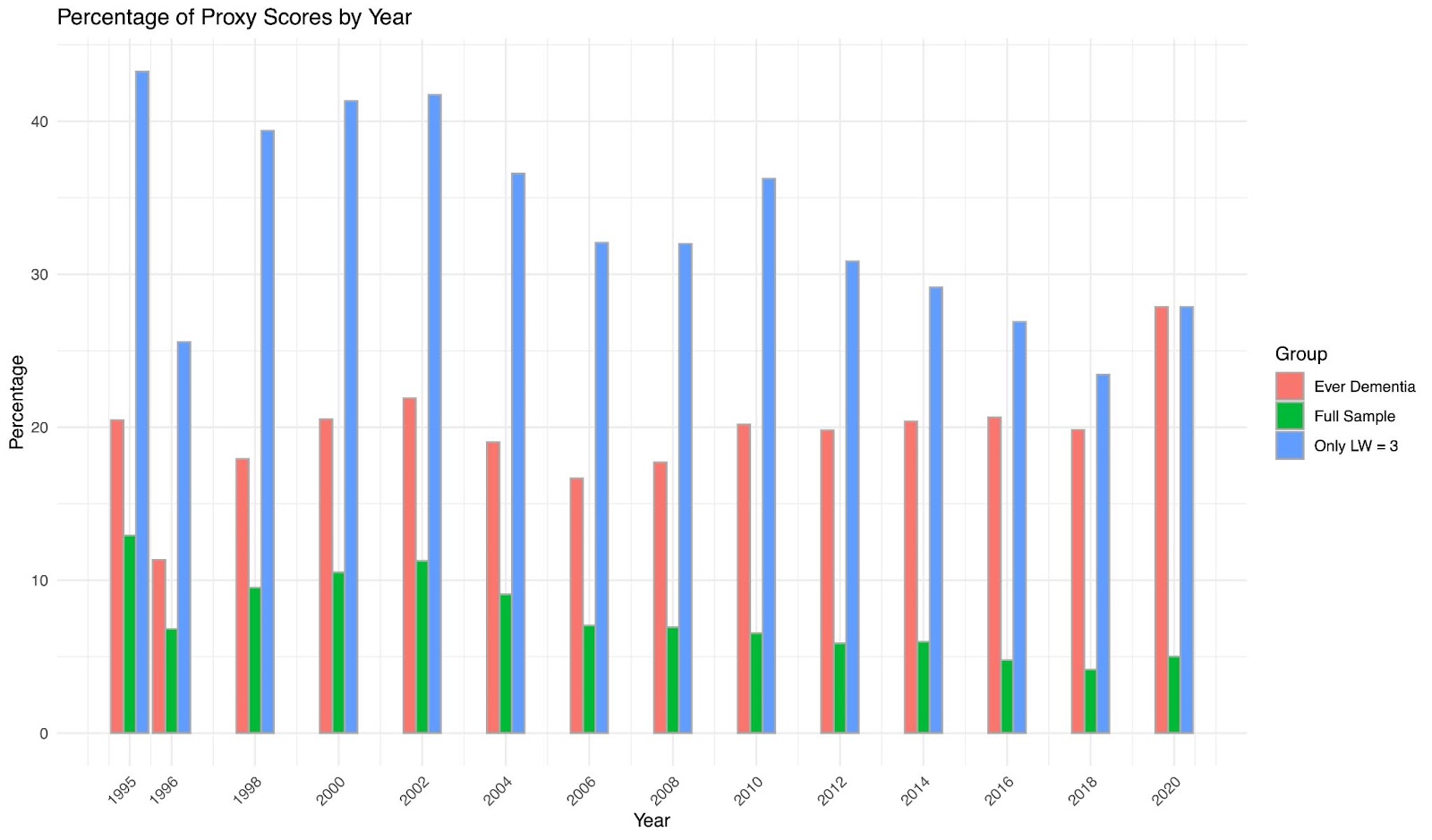

**Figure S2. Percentage of participants with Langa-Weir scores based on proxy data for each year of assessment in HRS.** Bars in red show the percentage of participants that are ever diagnosed with dementia that have a proxy score in that year of assessment, green bars show the percentage of participants with a proxy score per year for the full HRS sample, and blue bars show the percentage of participants that have a Langa-Weir score of 3 (indicating dementia) based on proxy data in a given assessment year. A large proportion of Langa-Weir scores of 3 at any given timepoint are based on proxy scores, indicating that many participants with dementia receive Langa-Weir scores based on proxy data.

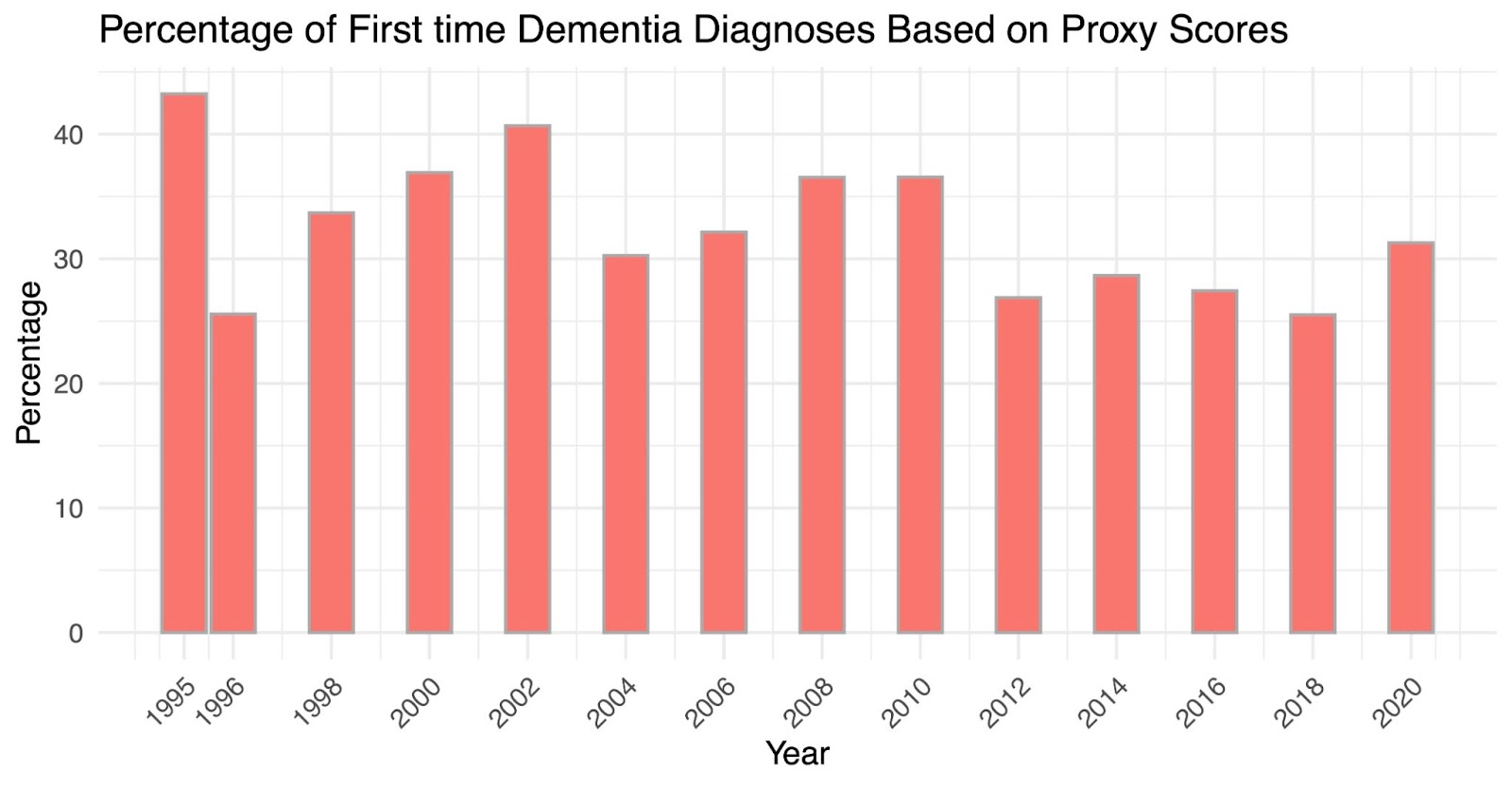

**Figure S3. Percentage of participants whose first Langa-Weir score of 3 is based on proxy data.** Bars show the percentage of participants in an assessment year that receive their first Langa-Weir score of 3 and that score is based on proxy data. For a given assessment year, between 30-40% of initial Langa-Weir scores of 3 are based on proxy data.

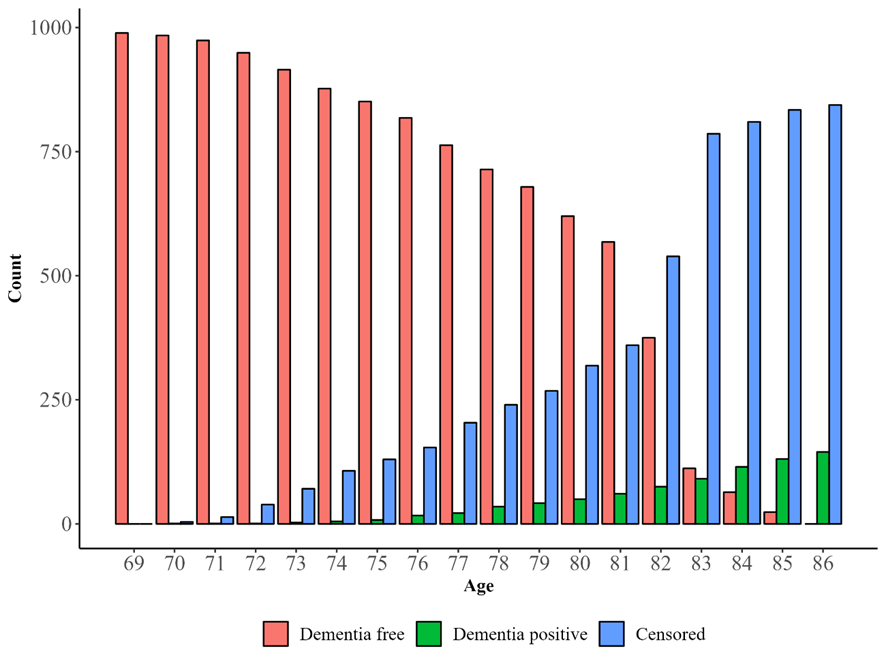

**Figure S4.** Absolute frequencies of dementia free participants, dementia positive participants, and censored cases across age groups in LBC1936 (N positive dementia diagnosis = 145; N total = 989).

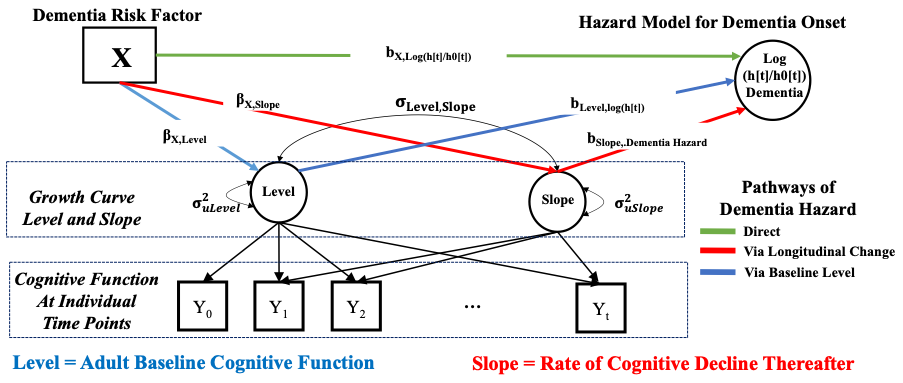

**Figure S5. Path diagram for the joint longitudinal-hazard model of cognitive levels, cognitive change, dementia hazard, and external predictors in HRS.** In the joint longitudinal-hazard model, cognitive level is regressed on the external predictor (X) (blue path), cognitive linear slope is regressed on the external predictor (red path) and the dementia hazard (log (h[t]) is regressed on the external predictor (green path).

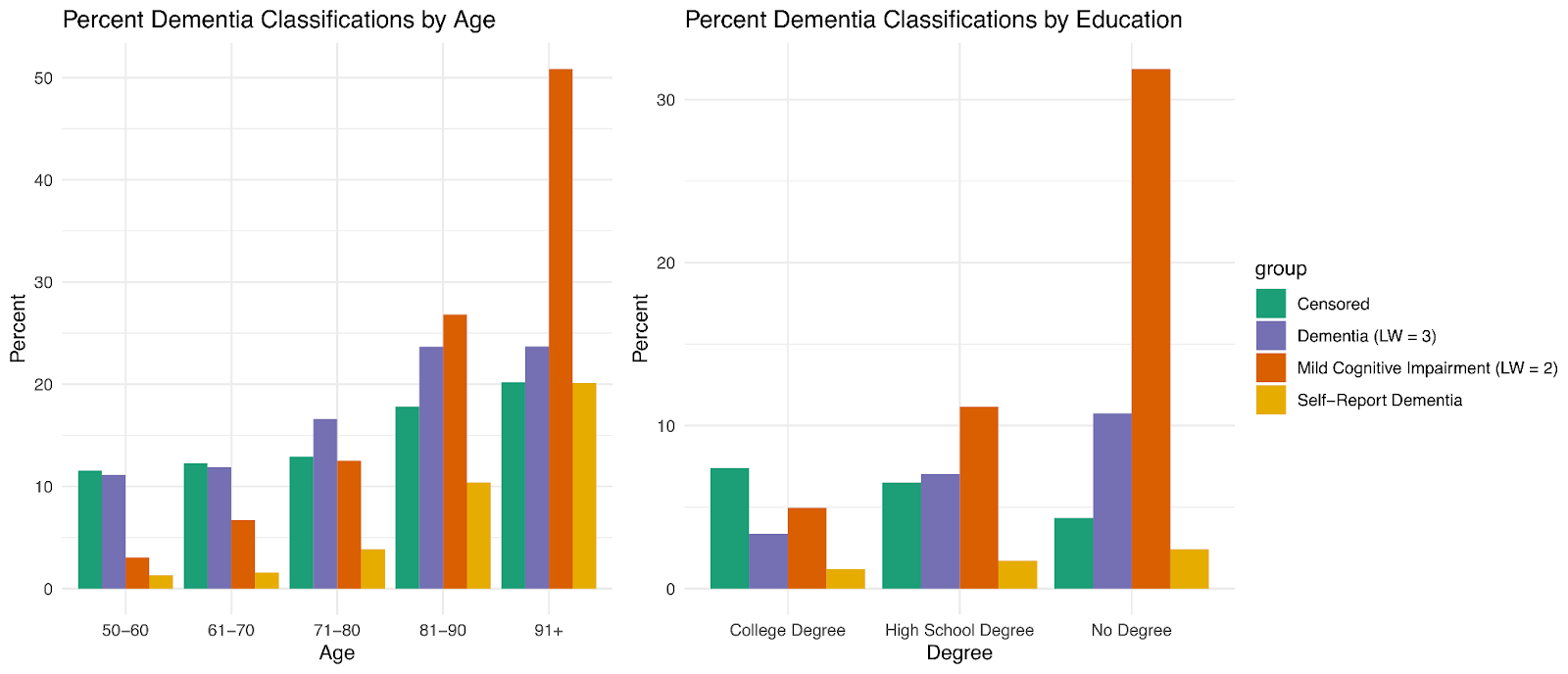

**Figure S6. Dementia prevalence rates for age groups and education groups within the sample.** Percent of participants that were right censored (green), diagnosed with dementia (purple), diagnosed with cognitive impairment (orange), or self-reported dementia (yellow) by age (left) and highest level of education (right). Dementia and cognitive impairment prevalence increases with older age and lower levels of educational attainment, consistent with prior literature. Self-reported dementia increases with age and with less education but is under-reported compared to Langa-Weir dementia scores.

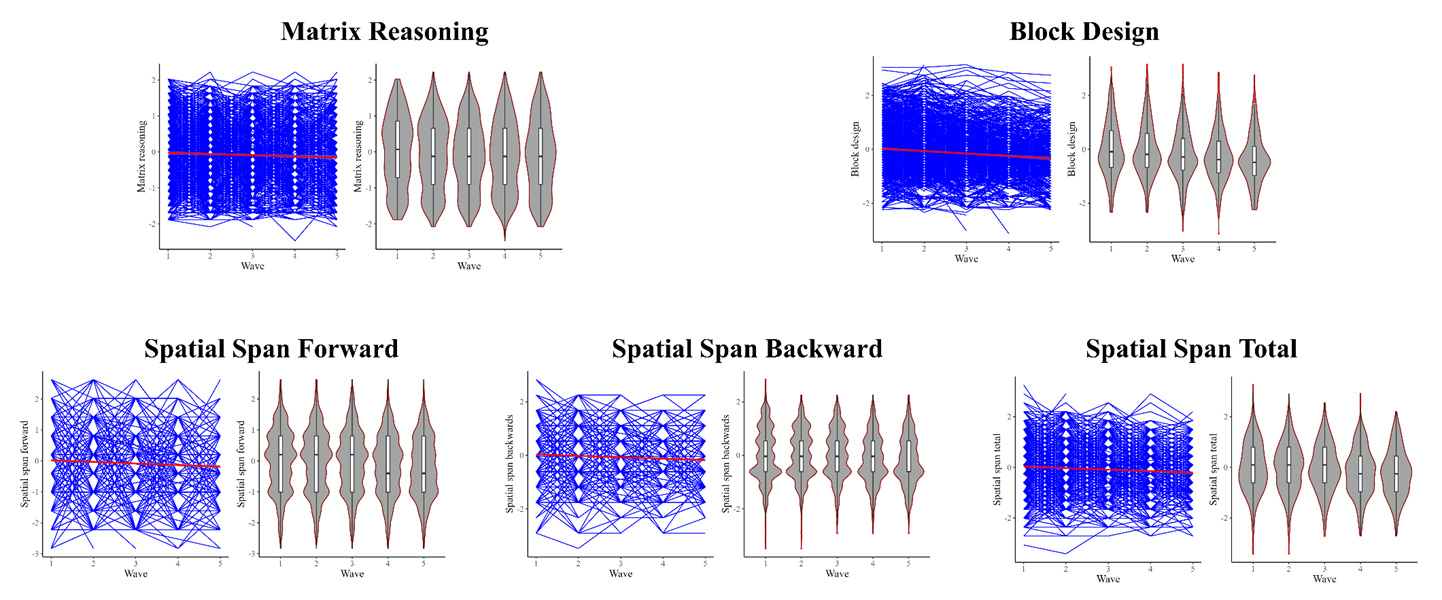

**Figure S7.** Distribution of visuospatial ability test scores across assessment waves in LBC1936.

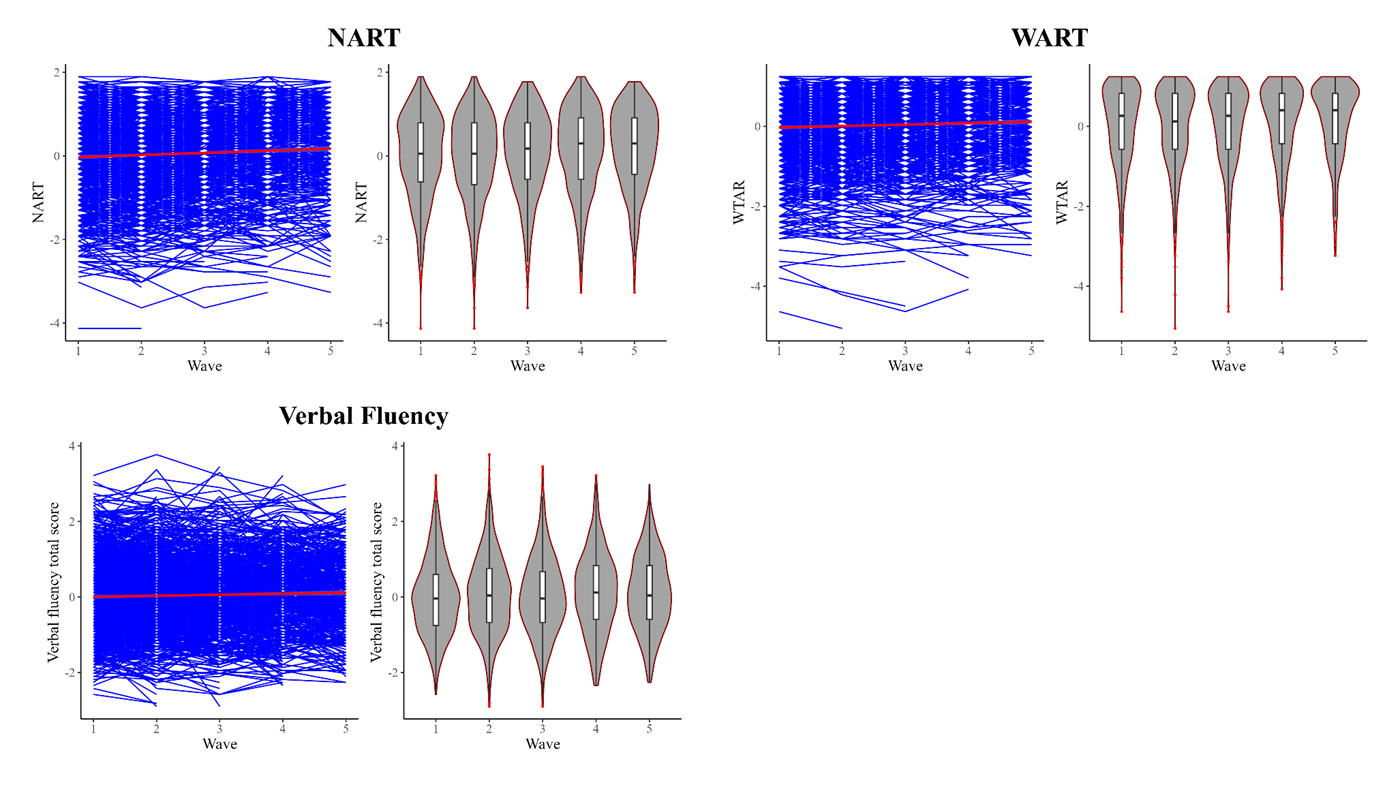

**Figure S8.** Distribution of crystallized knowledge test scores across assessment waves in LBC1936.

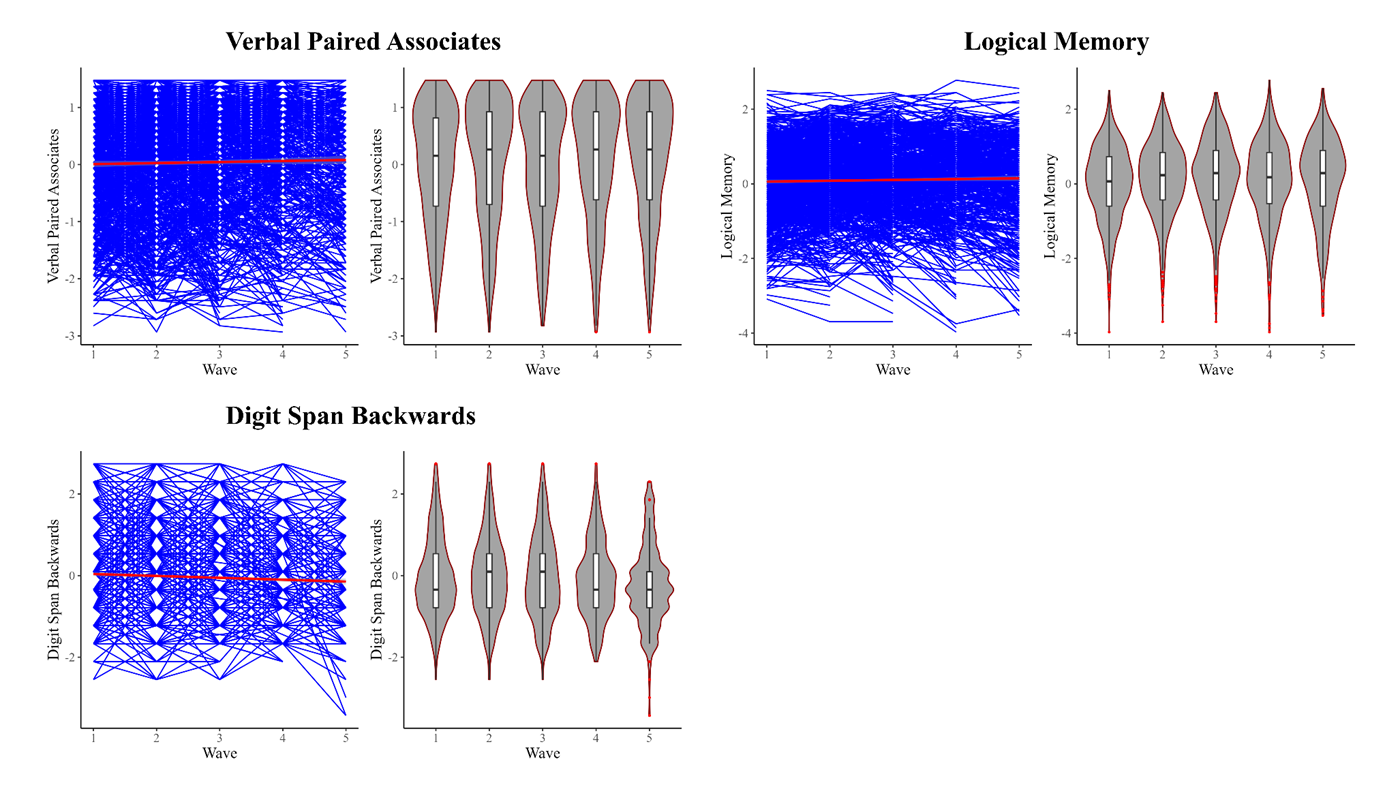

**Figure S9.** Distribution of verbal memory test scores across assessment waves in LBC1936.

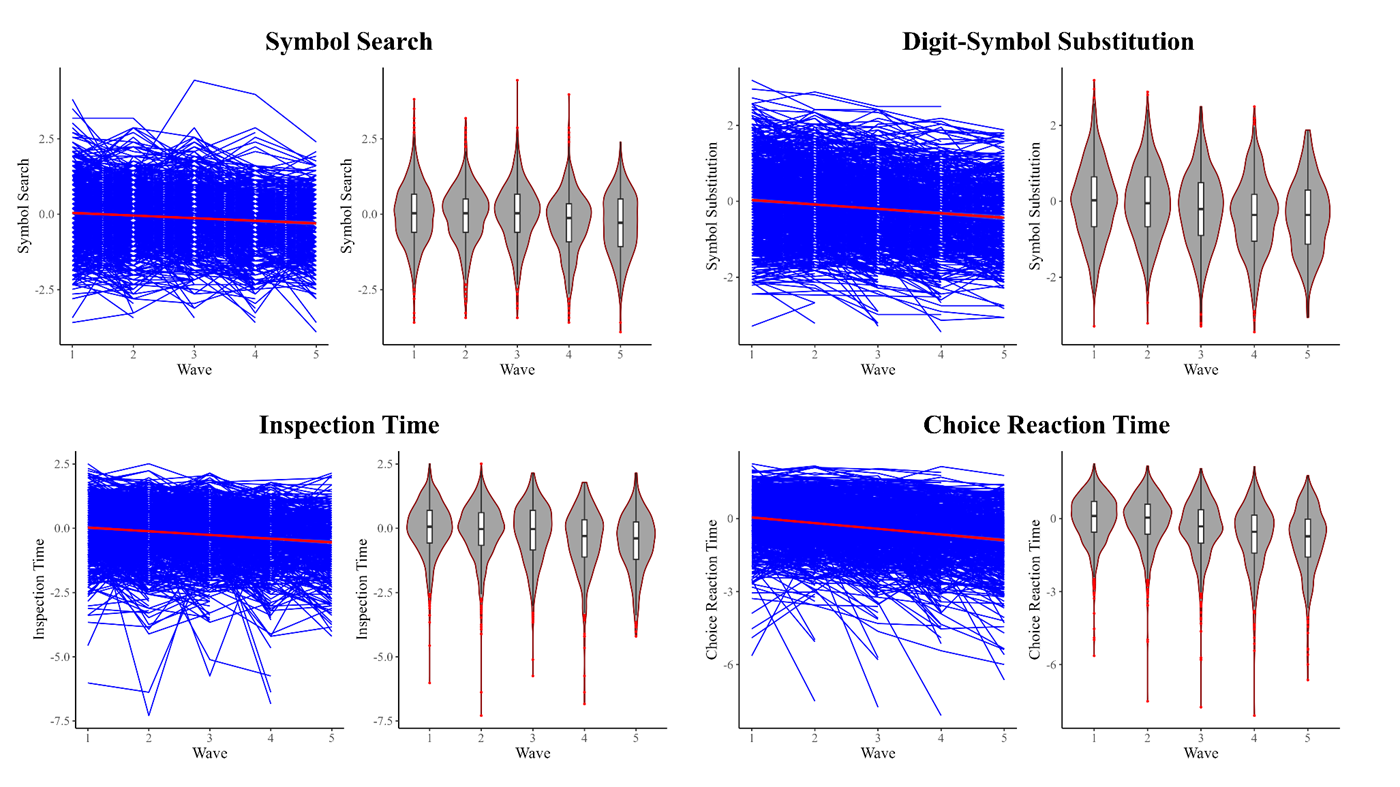

**Figure S10.** Distribution of processing speed test scores across assessment waves in LBC1936.

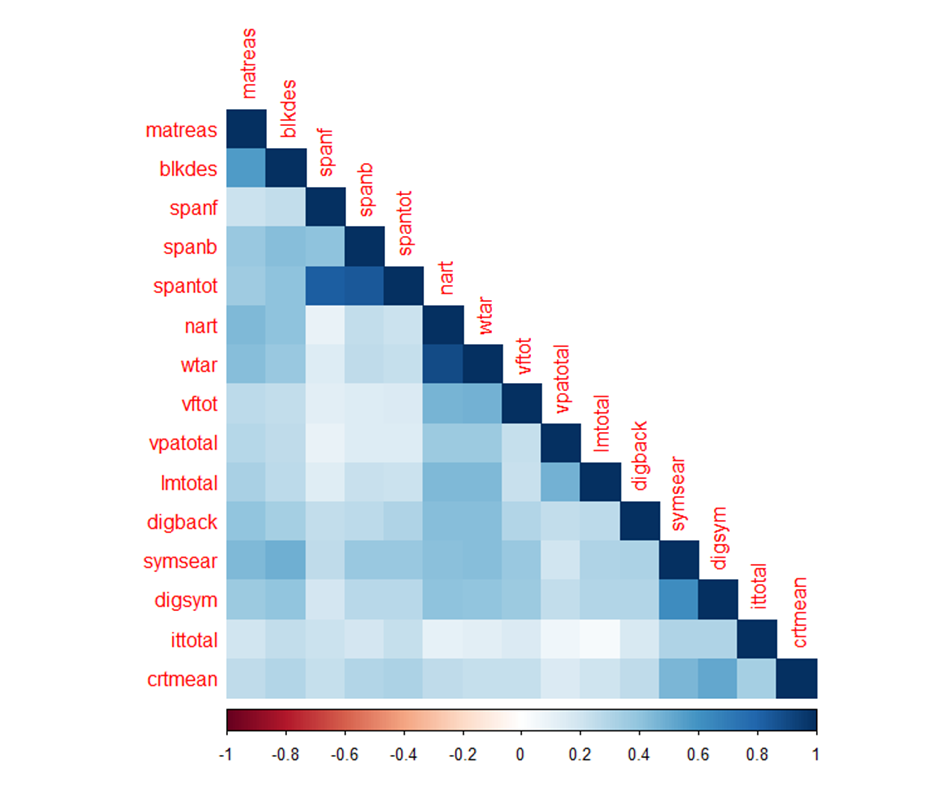

**Figure S11**. Heatmap of correlations across standardized cognitive tests scores at baseline assessment in LBC1936.
